# Household economic impact of road traffic injury versus routine emergencies in a low-income country

**DOI:** 10.1101/2020.09.29.20204081

**Authors:** Hani Mowafi, Brian Rice, Rashida Nambaziira, Gloria Nirere, Robert Wongoda, Matthew James, GECC Writing Group, Mark Bisanzo, Lori Post

## Abstract

**Introduction:** Road traffic injuries (RTIs) are increasing and have disproportionate impact on residents of low- and middle-income countries (LMICs) where 90% of deaths occur. RTIs are a leading cause of death for those aged 15 – 29 years with costs estimated to be up to 3% of GDP. Despite this fact, little primary research has been done on the household economic impact of these events.

**Methods:** From July to October 2016, 860 consecutive emergency department patients were enrolled and followed up at 6-8 weeks to assess the household financial impacts of these emergency presentations. At follow-up, patients were queried regarding health status, lost wages or schooling, household costs incurred due to their injury or illness, and assets sold.

**Results:** 860 patients were enrolled and 675 patients (78%) completed follow-up surveys. Of those, 661 had a confirmed reason for visit - 304 (45%) road traffic injuries, 357 (53%) other emergency presentations (non-RTI) - encompassing medical presentations and other types of injury. Data was missing for 14 patients (2%). More than 90% of RTI patients were working or in school prior to their injury. In the economically productive ages (15-44 years) RTI predominated (70%) vs non-RTI (39%). RTI patients were more likely to report residual major disability (28% RTI vs 21% non-RTI, p<0.03). All emergency patients reported difficulty paying for basic needs (food, housing and medical expenses). More than ⅓ of emergency patients reported having to sell assets in order to meet basic needs after their illness or injury. Despite similar hospital costs and fewer lost days of work for both patients and caregivers, the mean financial impact on households of RTI patients was 37% more than for non-RTI patients. These costs equaled between 6-16 weeks of income for patients based on their occupation type and median reported pre-hospitalization income.

**Discussion:** Ugandans emergency care patients suffered significant personal and household economic hardship. In addition to the need for policy and infrastructural changes to improve road safety, these findings highlight the need for basic emergency care systems to secure economic gains in vulnerable households and prevent medical impoverishment of marginal communities.

**Highlights:**

- Road traffic injuries (RTIs) and routine emergencies result in residual disability among the majority of patients in Uganda 6-8 weeks after their initial presentation with less than 20% reporting full recovery.
- The household financial impact from road traffic injuries is significant - ranging from the equivalent of 6 to 16 weeks of income - and is broadly felt across types of workers.
- Despite similar hospital costs and fewer lost days of work for both patients and caregivers, the mean financial impact on households of RTI patients was 37% more than for non-RTI patients, which included medical presentations and non-RTI injuries.
- All emergency patients reported difficulty paying for basic needs including food, housing and medical expenses 6-8 weeks post emergency unit encounter. More than ⅓ of emergency patients reported having to sell assets in order to meet basic needs after their illness or injury.
- Economic impacts extended beyond injured patients to family members, 79% of whom missed work or school to provide care to the patient.

## Introduction

The incidence of road traffic injury (RTI) is increasing globally with a disproportionate impact on residents of low- and middle-income countries (LMICs) where 90% of road traffic collisions global road traffic deaths occur (double the fatality rate of high-income countries).[1] Road traffic injuries are a leading cause of death for those aged 15 – 29 years and the 8th leading cause for all ages with costs estimated to be up to 3% GDP[1] and with lost potential growth of 7-22% GDP.[2] Severe road injuries or fatalities in this economically productive age group can result in catastrophic economic costs for households, especially poorer ones, in low-income countries.[3] Despite this fact, little primary research has been done on the economic impact of these events at the household level.

The number of RTIs has risen steadily, reaching 1.35 million in 2016, while deaths relative to the global population has remained constant.[1] The burden is particularly high in rapidly-developing low-income countries with increasing levels of motorization. There was a four-fold increase in road traffic fatalities in Uganda from 2011-2014 and the WHO estimates current annual traffic fatalities at 12,058.[4] There is a strong inverse association between RTI deaths and country income levels with RTI fatalities in low-income countries (LIC) being three times that in high-income countries (HIC).[1] While progress has been made towards improving road safety through policy and infrastructural improvements and through mitigating impacts of RTIs through improvements in pre-hospital and emergency care in many middle- and high-income countries, no reduction in road traffic deaths was seen in any low-income country between 2013 and 2016.[1] Vulnerable road users – including pedestrians and cyclists – are disproportionately affected and Africa has the highest proportion of mortality among vulnerable road users (44%).[1] Despite this fact, they have been largely ignored in the planning, design, and operation of roads.[1]

Many health systems in LMICs rely on out-of-pocket (OOP) expenditures for financing with such payments representing 39% of costs in low-income countries, 56% in lower-middle-income countries, and 30% in upper-middle-income countries (global average 18%).[5] These costs are borne by patients and their families in the immediate aftermath of their acute illness or injury. This has lead researchers to investigate the financial impact of these sudden shocks associated with acute and exacerbations of chronic conditions on households, especially the poor. Most published analyses have focused on infectious conditions such as malaria[6,7] or HIV[8–12] for which independent vertical programs exist in many countries for research and clinical care. Little attention has been paid to the financial impacts of more common conditions, like RTI, that have significant impacts on households in low-income communities. Furthermore, when a condition impacts an individual’s ability to continue in paid work or results in the death of a breadwinner, the entire household experiences negative economic consequences that are not adequately captured by measures such as catastrophic health expenditures (CHE) that fail to quantify lost or future earnings.[5]

Most analyses of economic impact of road traffic injuries are based on models that use aggregate health and economic data and surveys using the Human Capital Approach (HCA) or the Willingness to Pay (WTP) - each of which are prone to underestimates in LMICs where underlying data is limited. A recent systematic review found that it is likely that cost estimates are based on underestimates of total injuries and fatalities and that extrapolations of cost estimates across contexts – particularly from high-income countries to low-income countries – are severely limited and can’t replace contextual data collected primarily.[13]

RTI is projected to remain a top-10 cause of death globally for the next 20 years.[1] As worldwide efforts to improve standards of living and eradicate poverty continue, there must be awareness of the potential for emergent conditions, like RTI, to negatively impact these initiatives and hinder development efforts. The objective of this manuscript is to quantify the household economic effects of RTI vs non-RTI emergency presentations in Masaka, Uganda.

## Methods

From July to October 2016 a prospective study was conducted at Masaka Regional Referral Hospital in Masaka, Uganda that enrolled 860 consecutive patients presenting to the emergency unit to assess the financial impacts of these emergency presentations on patients and their households. Masaka District Hospital is a public hospital operated by the Ugandan Ministry of Health and is the regional referral hospital for 8 districts (Kalangala, Lyantonde, Masaka, Sembaule, Kalungu, Lwengo, Bukomansimbi, and Rakai). The hospital averages 65 daily admissions.

Patients were enrolled by trained research assistants and consented for a follow-up survey at 6-8 weeks after the date of their presentation to assess the post-hospitalization impacts at the household level. Patients at this facility are routinely contacted by hospital staff 3 days after their emergency visit to assess their clinical outcomes. At the time of presentation, enrolled patients provided a phone number (their own, that of a family member or neighbor) and follow-up calls were made to the provided number between 6-8 weeks post initial presentation.

At follow-up, patients were queried regarding their health status, costs their household incurred as a result of illness or injury, any lost wages or time at school for the patient and/or caregivers, whether they were forced to sell any assets, and whether they had difficulty meeting basic needs including purchasing food, housing, and medicine. (Appendix 1 – Survey instrument). Survey responses were collated in an online survey tool (Qualtrics, 2016, version Aug-Oct). Descriptive statistics were calculated, and comparisons were made between RTI patients and non-RTI patients presenting to the Masaka hospital emergency unit.

The study was approved by the ethical review boards of Makerere University and Yale University.

## Results

During the study period, 860 patients were enrolled, and follow-up surveys were completed on 675 patients (78%) with 185 patients lost to follow-up (22%). Of the 675 patients there were 304 RTI patients (45%), 357 non-RTI patients (53%) and 14 patients missing chief complaints (2%). Of those lost to follow-up, 157 could not be reached after 3 attempts in 10 days and 28 where the wrong phone number had been given/recorded. The comparison of those on whom follow-up was obtained and those lost to follow-up is included in Appendix 2.

### Patient Characteristics

The characteristics of patients who completed the follow-up survey are presented in Table 1. RTI patients were more likely to be male, employed and young adults than non-RTI patients. Overall, more than 90% of the RTI patients were working or in school prior to their injury event. Non-RTI patients were more likely to be from the extremes of age, slightly more likely to be female, and were employed at a lower rate than RTI patients.

**Table 1:** Characteristics Emergency Department Patients

|  | RTI |  | Not RTI |  | p-value |
| --- | --- | --- | --- | --- | --- |
|  | n | % | n | % |  |
|  | 304 | 46.0% | 357 | 54.0% |  |
| Male | 211 | 52.9% | 188 | 47.1% | **<0.001** |
| Age (years) |  |  |  |  |  |
| Under 5 | 19 | 6.3% | 69 | 19.3% |  |
| 05-14 | 29 | 9.5% | 42 | 11.8% |  |
| 15-44 | 213 | 70.1% | 139 | 38.9% |  |
| 45-64 | 34 | 11.2% | 55 | 15.4% | **<0.001** |
| >= 65 | 3 | 1.0% | 44 | 12.3% |  |
| UNK | 6 | 2.0% | 8 | 2.2% |  |
| Student? | 47 | 15.5% | 53 | 14.8% | 0.83 |
| Employed? | 229 | 75.3% | 180 | 50.4% | **<0.001** |
| Type of Work (if employed) |  |  |  |  |  |
| Manual | 63 | 27.5% | 81 | 45.0% |  |
| Unskilled | 57 | 24.9% | 21 | 11.7% | **<0.001** |
| Skilled | 40 | 17.5% | 21 | 11.7% |  |
| Professional | 69 | 30.1% | 57 | 31.7% |  |

### Health Status

RTI and non-RTI patients were equally likely to be fully recovered or report minor disability at follow-up. RTI patients were significantly more likely to report major disability preventing return to work or school at follow up (28%) vs non-RTI patients (21%) and this finding held true even for those who were not admitted and discharged directly from the emergency unit (Table 2). There were more fatalities recorded among non-RTI patients than RTI patients who survived to hospital presentation.

**Table 2:** Morbidity and Morality Outcomes

|  | RTI |  | Not RTI |  | p-value |
| --- | --- | --- | --- | --- | --- |
|  | n | % | n | % |  |
|  | 304 | 46.0% | 357 | 54.0% |  |
| Full Recovery | 58 | 19.1% | 74 | 20.7% | 0.6 |
| Minor Disability | 148 | 48.7% | 155 | 43.4% | 0.18 |
| Major Disability | 86 | 28.3% | 75 | 21.0% | 0.03 |
| Death* | 7 | 2.3% | 40 | 11.2% | <0.001 |

\*Includes only patients that survived transport to hospital. Excludes prehospital deaths.

### Caregiver impact

Among RTI patients, 241 (79%) reported a family member missed days of work or school to care for them and the mean number of days of work missed by caregivers was 18.8 days. Non-RTI patients reported a similar percentage of caregivers missing work (197, 88%) and a similar mean of 21.7 days (Table 3).

**Table 3:**
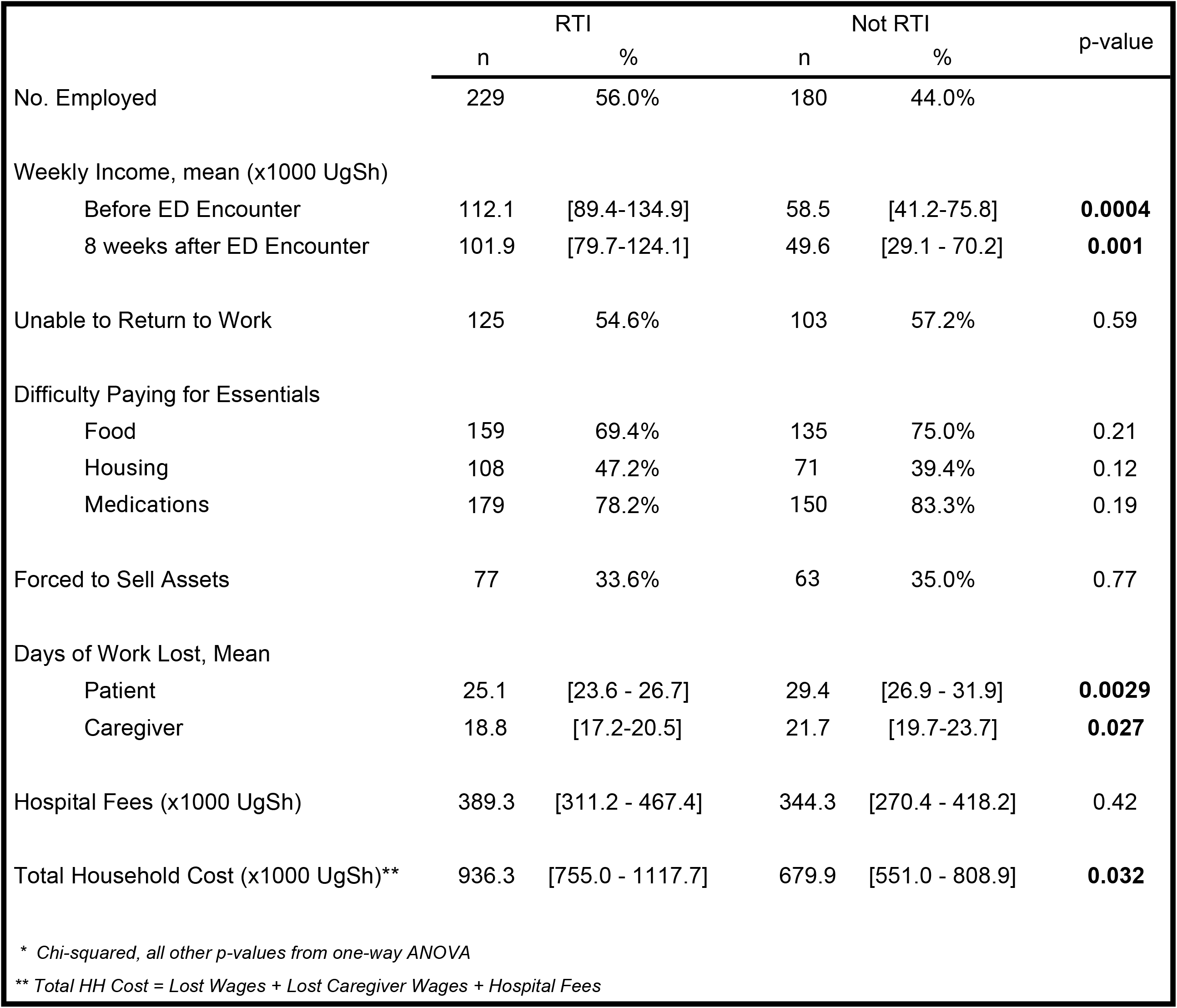
Financial Impact on Working Patients

|  | RTI |  | Not RTI |  | p-value |
| --- | --- | --- | --- | --- | --- |
|  | n | % | n | % |  |
| No. Employed | 229 | 56.0% | 180 | 44.0% |  |
| Weekly Income, mean (x1000 UgSh) |  |  |  |  |  |
| Before ED Encounter | 112.1 | [89.4-134.9] | 58.5 | [41.2-75.8] | **0.0004** |
| 8 weeks after ED Encounter | 101.9 | [79.7-124.1] | 49.6 | [29.1 - 70.2] | **0.001** |
| Unable to Return to Work | 125 | 54.6% | 103 | 57.2% | 0.59 |
| Difficulty Paying for Essentials |  |  |  |  |  |
| Food | 159 | 69.4% | 135 | 75.0% | 0.21 |
| Housing | 108 | 47.2% | 71 | 39.4% | 0.12 |
| Medications | 179 | 78.2% | 150 | 83.3% | 0.19 |
| Forced to Sell Assets | 77 | 33.6% | 63 | 35.0% | 0.77 |
| Days of Work Lost, Mean |  |  |  |  |  |
| Patient | 25.1 | [23.6 - 26.7] | 29.4 | [26.9 - 31.9] | **0.0029** |
| Caregiver | 18.8 | [17.2-20.5] | 21.7 | [19.7-23.7] | **0.027** |
| Hospital Fees (x1000 UgSh) | 389.3 | [311.2 - 467.4] | 344.3 | [270.4 - 418.2] | 0.42 |
| Total Household Cost (x1000 UgSh)** | 936.3 | [755.0 - 1117.7] | 679.9 | [551.0 - 808.9] | **0.032** |
| * Chi-squared, all other p-values from one-way ANOVA |  |  |  |  |  |
| ** Total HH Cost = Lost Wages + Lost Caregiver Wages + Hospital Fees |  |  |  |  |  |

### Lost Wages

Patients and family members missed a substantial amount of work after their emergency injury or illness. All patients averaged nearly a month of missed work (RTI 25.1 days vs non-RTI 29.4) (Table 3). Among all occupation groups, caregivers missed 2 - 3 weeks of work. There was a substantial percentage drop in income across each occupational group for RTI patients (Table 4).

**Table 4:**
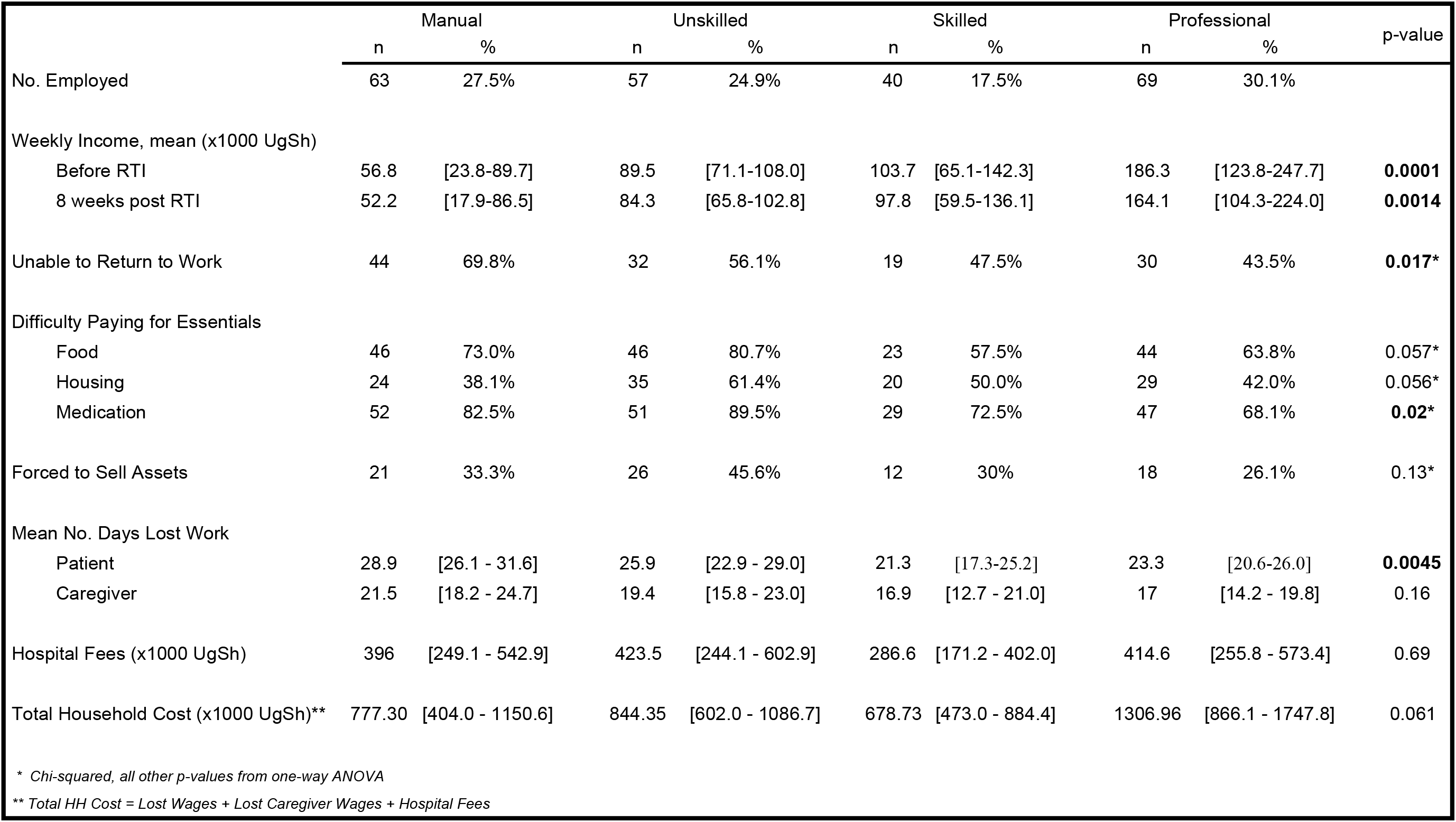
Subgroup Analysis of Financial Impact of RTI by Type of Work

|  | Manual |  | Unskilled |  | Skilled |  | Professional |  | p-value |
| --- | --- | --- | --- | --- | --- | --- | --- | --- | --- |
|  | n | % | n | % | n | % | n | % |  |
| No. Employed | 63 | 27.5% | 57 | 24.9% | 40 | 17.5% | 69 | 30.1% |  |
| Weekly Income, mean (x1000 UgSh) |  |  |  |  |  |  |  |  |  |
| Before RTI | 56.8 | [23.8-89.7] | 89.5 | [71.1-108.0] | 103.7 | [65.1-142.3] | 186.3 | [123.8-247.7] | **0.0001** |
| 8 weeks post RTI | 52.2 | [17.9-86.5] | 84.3 | [65.8-102.8] | 97.8 | [59.5-136.1] | 164.1 | [104.3-224.0] | **0.0014** |
| Unable to Return to Work | 44 | 69.8% | 32 | 56.1% | 19 | 47.5% | 30 | 43.5% | **0.017\*** |
| Difficulty Paying for Essentials |  |  |  |  |  |  |  |  |  |
| Food | 46 | 73.0% | 46 | 80.7% | 23 | 57.5% | 44 | 63.8% | 0.057* |
| Housing | 24 | 38.1% | 35 | 61.4% | 20 | 50.0% | 29 | 42.0% | 0.056* |
| Medication | 52 | 82.5% | 51 | 89.5% | 29 | 72.5% | 47 | 68.1% | **0.02\*** |
| Forced to Sell Assets | 21 | 33.3% | 26 | 45.6% | 12 | 30% | 18 | 26.1% | 0.13* |
| Mean No. Days Lost Work |  |  |  |  |  |  |  |  |  |
| Patient | 28.9 | [26.1 - 31.6] | 25.9 | [22.9 - 29.0] | 21.3 | [17.3-25.2] | 23.3 | [20.6-26.0] | **0.0045** |
| Caregiver | 21.5 | [18.2 - 24.7] | 19.4 | [15.8 - 23.0] | 16.9 | [12.7 - 21.0] | 17 | [14.2 - 19.8] | 0.16 |
| Hospital Fees (x1000 UgSh) | 396 | [249.1 - 542.9] | 423.5 | [244.1 - 602.9] | 286.6 | [171.2 - 402.0] | 414.6 | [255.8 - 573.4] | 0.69 |
| Total Household Cost (x1000 UgSh)** | 777.30 | [404.0 - 1150.6] | 844.35 | [602.0 - 1086.7] | 678.73 | [473.0 - 884.4] | 1306.96 | [866.1 - 1747.8] | 0.061 |
| * Chi-squared, all other p-values from one-way ANOVA |  |  |  |  |  |  |  |  |  |
| ** Total HH Cost = Lost Wages + Lost Caregiver Wages + Hospital Fees |  |  |  |  |  |  |  |  |  |

### Economic impact

A majority of patients in both groups reported difficulty paying for such essential items as food, shelter and medicine as a result of their condition at 6 – 8 week follow-up (Table 3). Roughly 30% of patients reported having to sell assets, most frequently in the form of livestock, in order to help pay for medical expenses.

While both RTI and non-RTI patients reported a drop in their weekly earnings at follow up, RTI patients reported a mean income that was significantly higher than non-RTI both before and after their emergency encounter (Table 3). When patients were stratified by occupational group (Appendix 3) the total economic impact on households (sum of out-of-pocket (OOP) and lost wages (LW) for patients and caregivers) was significant across all occupational groups but greatest for RTI patients who worked as professionals when measured in absolute terms (Figure 1, 2). However, when total household economic impact was converted into equivalent weeks of wages the impact of both RTI and non-RTI emergency presentations was most acutely felt by manual laborers (RTI ∼ 14 weeks wages vs non-RTI ∼ 22 weeks wages) (Figure 1, 2).

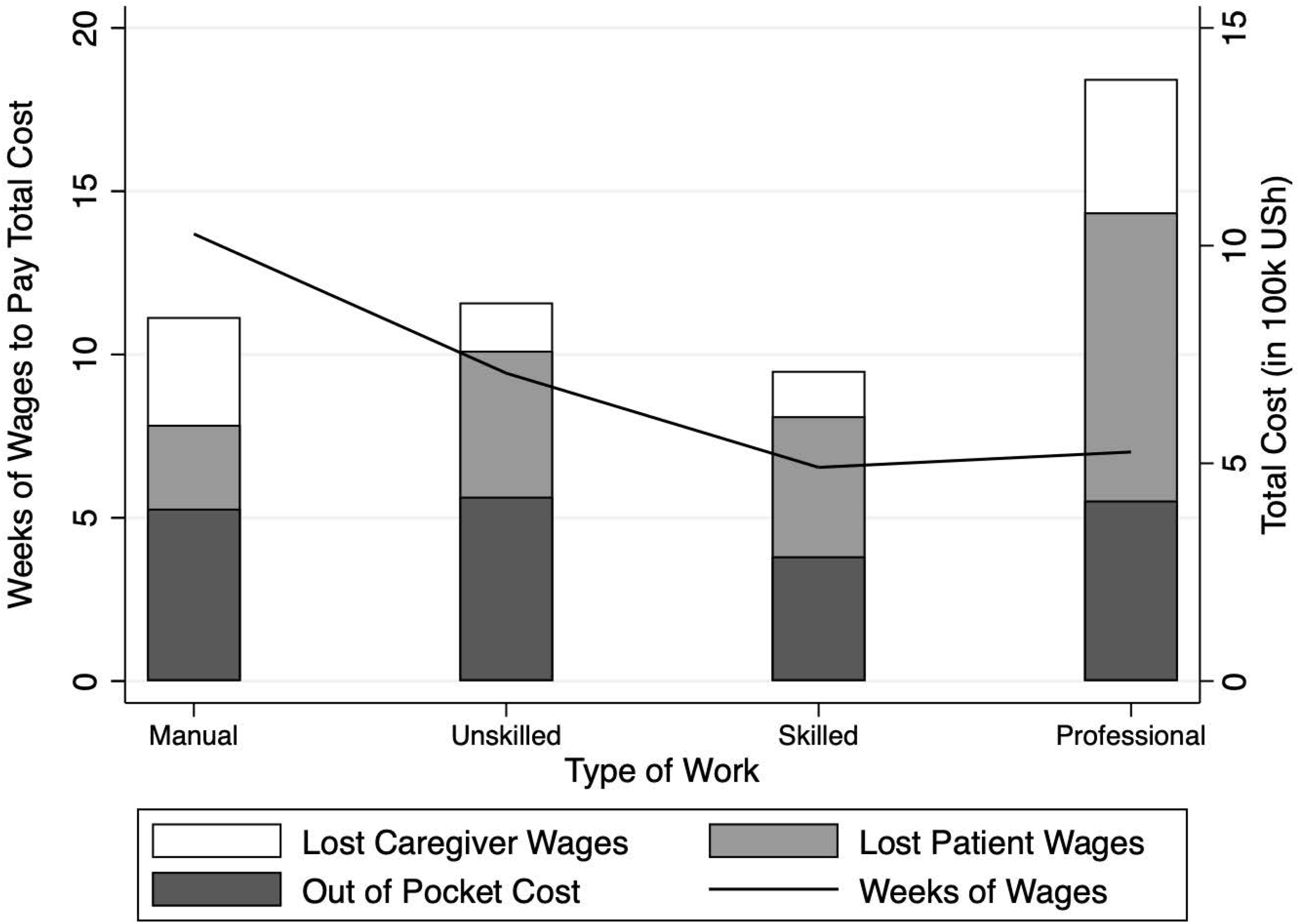

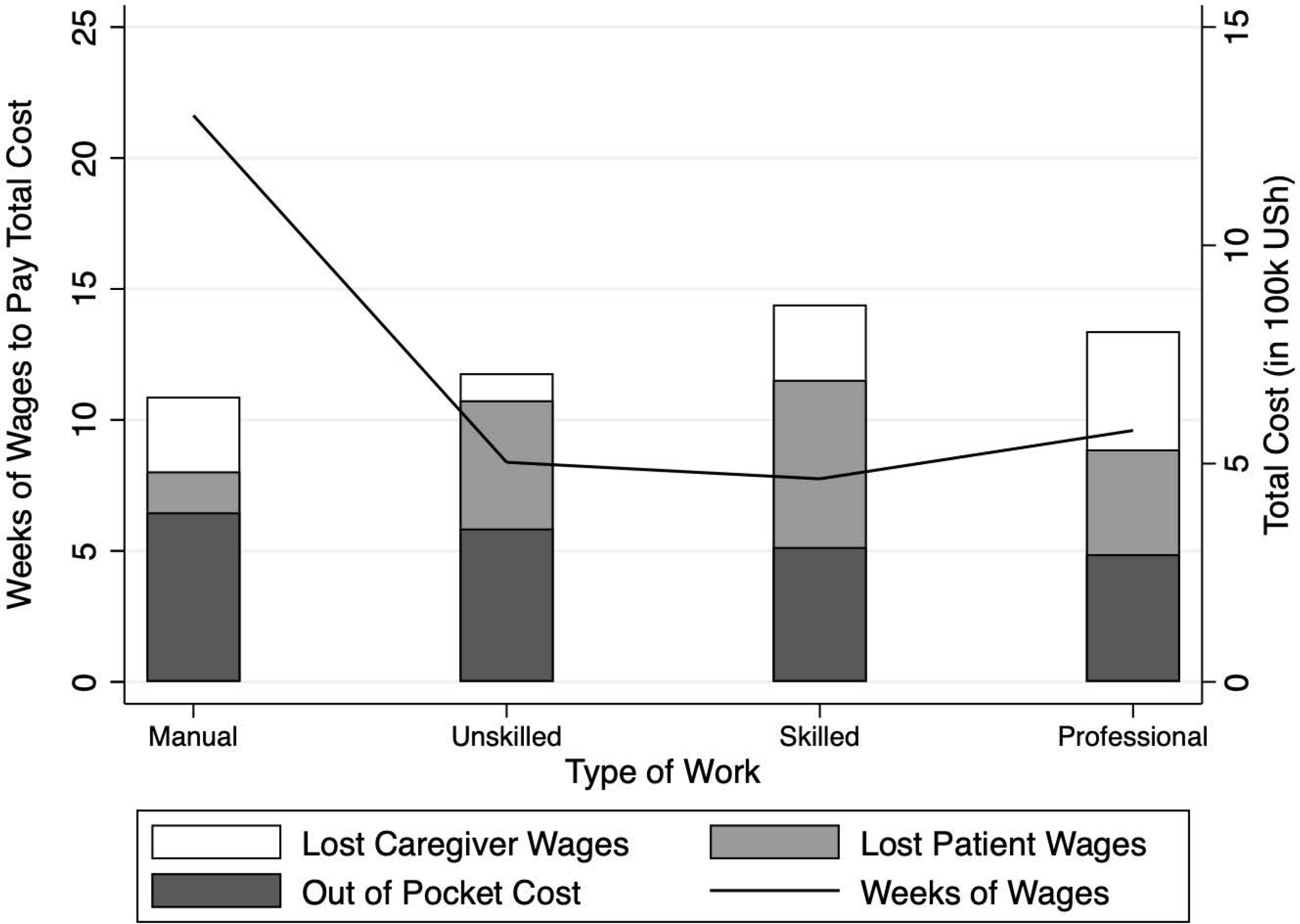

### Limitations

There were several limitations to this study that should be noted. First, since our study enrolled patients at the emergency unit, it could not capture deaths of those who did not survive to hospital presentation. Events with high potential costs such as sudden death of a breadwinner due to RTI would be absent in this dataset resulting in a likely under- estimate of total household economic impact. Moreover, prehospital costs that could be charged to the patient were not captured such as on-scene care, prehospital transport as well as property damage that may be borne by a household (such as damage or loss of a household vehicle) were also not captured as part of this study.

In addition, this survey was conducted at a single site and the period during which the study was conducted likely did not capture seasonal variability in workers income. These limitations could limit the generalizability to other patients in the country or region. Further, many patients were unemployed and reported zero income despite reporting an occupational group with higher mean income (e.g. professionals). This served to minimize differences between groups. Lastly, there were 185 (22%) patients that were lost to follow-up with only 31 (3.7%) due to an incorrect number. While every attempt was made to follow up repeatedly with these patients, the inability to document their outcomes limits the ability of this study to estimate the true economic impact on households as these households may have suffered catastrophic losses from the death of the injured family member that would not be documented here.

## Discussion

This study demonstrated that Ugandans who suffered RTIs as well as those who presented for other emergency complaints suffered significant morbidity and mortality as well as personal and household economic hardship. The mortality rate of RTIs was significantly lower than for other emergencies (2.3% vs 11.2%, p<0.001). As mentioned in limitations above, this reported RTI mortality rate substantially underestimates the true mortality associated with RTIs as this study only includes RTI patients that *survived transport to the hospital*. Comparisons of facility-reported RTI deaths with nationally-reported RTI deaths suggest that only 10-25% of RTI fatalities survive transport to eventually die within a facility, thus up to 90% of road fatalities would not be captured in this study.[1,14] Controlling for this reality was outside of the scope of our study. However, even recognizing that the most severely injured patients died on scene, our results showed that RTI patients that did survive transport to a facility experienced significantly higher rates of major disability than did non-RTI emergencies (28.3% vs. 21.0%, p=0.03).

The majority of households reported difficulty paying for essential items including food, shelter and medical costs. Further, those who previously worked reported continued disability and inability to return to work eight weeks after their emergency event. Most households reported that family members missed weeks of work and school to provide care to their ill or injured relatives. Total costs of RTI patients were significantly higher than non-RTI patients even when these costs reflected only a portion of the expected totals.

Previous work by Ross Silcock and the Transport Research Laboratory suggests that the human capital approach (HCA) is preferred to the willingness-to-pay (WTP) method for estimating total costs of RTIs in LMICs since WTP requires completion of lengthy questionnaires and relies on valuation of perceive risk from hypothetical events. In using HCA it is suggested that indirect costs associated with grief, suffering, residual disability and the sudden financial drain associated with premature household deaths may be estimated as a percentage of direct costs using 28% of total direct costs (TDC) for a fatal crash, 50% TDC for a serious crash, 8% TDC for a slight crash and 0% for property damage only.[15] While nearly 30% of RTI presentations reported residual disability or death, we did not attempt to quantify this additional cost because: a) we lacked the prehospital and property damage costs, b) reported residual disability at follow up is a proxy for severity of the accident but details of the RTIs were not available, and c) the inability to capture prehospital deaths precluded an accurate estimation of this cost.

Similar disabilities and financial impacts to those described above were seen in a study of patients in Malawi who suffered traumatic lower extremity injury.[16] Patients described difficulty with activities of daily living leading to food insecurity, the sale of assets, and missed school- and work-days by family members. Our study expands on these findings by also quantifying the drops in household income as well as the total household impact in terms of weeks of labor needed to cover the sudden, unexpected costs.

Most of the patients in this study were manual and unskilled laborers in a rural area and some evidence suggests this population is most vulnerable to suffering economic hardship after a road traffic accident[2]. Tayeb et al interviewed Sudanese households who had experienced injuries in the previous year.[17] They found that two-thirds of those injured had persistent disability related to an extremity injury. They also found that 15% of people in the lowest socioeconomic status lost their job because of their injury compared to just 5% in the highest quintile. It is reasonable to infer that the loss of function of an arm or leg would have a more significant impact on the ability to perform manual labor than to sell merchandise or participate in office-based work.

While our study focuses on the loss of earning potential over the short-term of 8 weeks, it is highly likely that economic disadvantages persisted well beyond this time frame. In a Korean survey, 71% of people who were disabled in an RTI experienced job loss, and on average took more than 3 years to obtain another form of employment.[18] This effect is likely greater in low-income countries like Uganda where it is estimated that 72% of the population is employed in agriculture.[19]

This study demonstrates the adverse impact of RTIs on development efforts in a low-income country, where not just individuals but entire households can be tipped into poverty by these sudden events. A variety of best practices exist for reducing the impact of RTIs including vehicle safety improvements (e.g. enforcement of speed laws; use of safety belts, helmets, and child restraints; as well as infrastructure like speed bumps and sidewalks). Also important are investments in emergency care systems to limit the negative impacts of these events when they occur. While the number of road traffic collisions continues to climb in high-income countries, the presence of emergency care systems to identify, transport, and resuscitate RTI patients can be the critical difference between a temporary event and one that has catastrophic impacts for households and communities.[1]

Rural road building is a major tool employed by donors and development agencies for poverty reduction in LMICs. While improved roads carrying more vehicles at higher speeds result can provide aggregate economic benefits to communities, this study demonstrates that they also produce negative externalities that severely negatively impact communities who are the specific targets of poverty reduction programs. In order to preserve economic gains and mitigate negative externalities we argue that such projects should not only be paired with investments in road safety infrastructure but also, importantly, in effective emergency care systems including timely transport to health facilities, necessary equipment and training of personnel in essential emergency care to provide life-saving care to RTI patients as well as to the wider community targeted for poverty reduction. At present, these are routinely lacking in LMIC communities, but there is hope. Pilot projects like the one to develop emergency medical services alongside the World Bank funded Southern Africa Trade and Transport Facilitation Program that is building high-speed motorways through Malawi and Tanzania is helping to quantify the impact that such investments can have on RTI mortality rates in these communities. Such analyses promise to give policy-makers for the first-time the ability to assess the cost-effectiveness of such interventions to save lives and preserve economic gains of development programs. As a result of this and other successful recent pilots in 2019-20, the World Bank will now require a global Road Safety Screening and Appraisal Tool (RSSAT) be used for all World Bank transport projects (and recommended for other operations with road safety impacts, such as urban or agricultural projects) to avoid or minimize road safety risks and impacts.[20] Such efforts when combined with empirical evidence gathered at the household level in LMICs to inform the analyses can improve development programs and help prevent communities on the economic margins from falling back into poverty.

## Data Availability

Data is available upon request at DRYAD Data server

https://doi.org/10.5061/dryad.tdz08kpvt

## Acknowledgements

The authors are grateful to the U.S. Agency for International Development (USAID) and the U.S. Department of Agriculture (USDA) for their funding of this work through the Feed the Future Program. The contents of the article are the responsibility of the authors and do not necessarily represent the views of the agencies or of the U.S. government.

The Authors declare that they have no conflicts of interest.

## Default Question Block

Please indicate who is recording the data

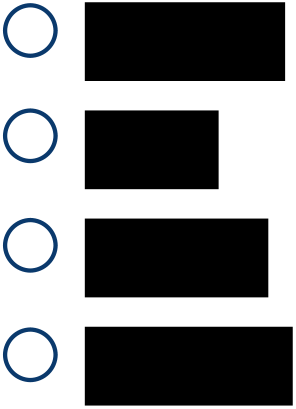

Good day. I hope you are well.

I am researcher calling from the Global Emergency Care Collaborative working at Masaka Hospital. We are conducting a research project along with Yale University in the United States under the approval of Makerere University in Kampala on the effects of emergency injury and illness in Uganda. We have a few short questions we would like to ask you regarding how you have been impacted by your recent illness or injury.

The information you may provide here is kept ***completely confidential and will not be shared with anyone other than the research team***. It will also be ***anonymous and your name and other identifiable information will not be reported*** along with your response.

You are under no obligation to participate and you may withdraw or stop participating at any time.

However, your assistance is greatly appreciated and will be used to improve emergency health services and public health planning for all Ugandans.

> *\*\*\*If they have not been previously consented, then please obtain verbal consent here. Preferable to either record that consent or have witnessed by another person and both initial if consent done on the phone\*\*\**
>
> *\*\*\* All the questions are geared towards the patient. For family members answering on behalf of the patient (e*.*g. in case of children or deceased patient) make sure they understand to whom the question is referring\*\*\**

Please record study ID # (assigned by research team, separate from the medical record number)

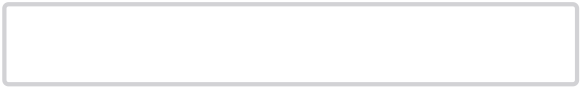

Please record last 4 digits of phone number

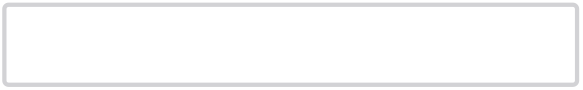

Type of consent

◯ Previous written
◯ Verbal in emergency department
◯ Verbal on phone
◯ Refuses consent

Follow-up status

◯ Successful Follow-up
◯ Lost to follow up, unable to reach after 3 attempts over 10 days
◯ Lost to follow up, wrong number

Who is the person being interviewed?

◯ Patient
◯ Parent or family member of patient on their behalf
◯ Other

If verbal consent, please record research staff names who are witnesses

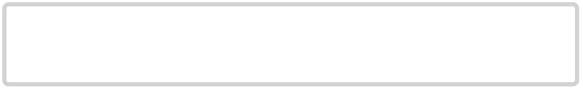

Your visit to the emergency department was for…

◯ Road Traffic Accident
◯ Another injury
◯ A medical condition not related to an injury

How would you report your current health status?

◯ Fully Recovered
◯ Partially recovered, continued minor health problems - able to work / return to school / return to usual activities
◯ Partially recovered, continued major health problems - unable to work / return to school / return to usual activities
◯ My health is the same as when I was first ill or injured
◯ My health status is worse than when I came to the emergency department
◯ Expired

Prior to your recent illness did you…

◯ Work (including working at home or on farm)
◯ Attend school
◯ Neither

Prior to your recent illness or injury, what type of work, if any, did you engage in? (Please list working at home as an option - if applicable)

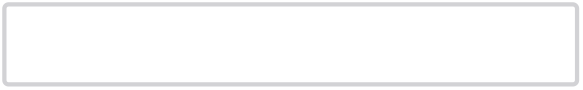

BEFORE YOUR RECENT ILLNESS / INJURY what was your ESTIMATED weekly income?

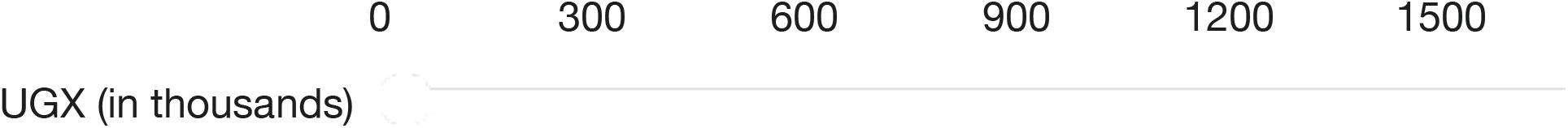

Please estimate the number of days that YOU have missed from WORK due to your recent illness or injury

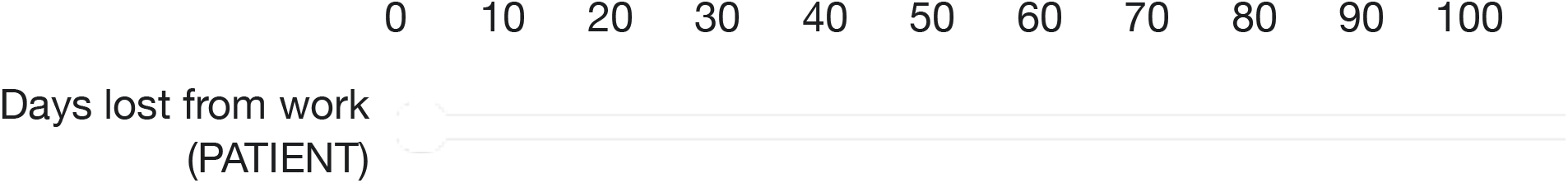

Please estimate the number of days that YOU have missed from SCHOOL due to your recent illness or injury

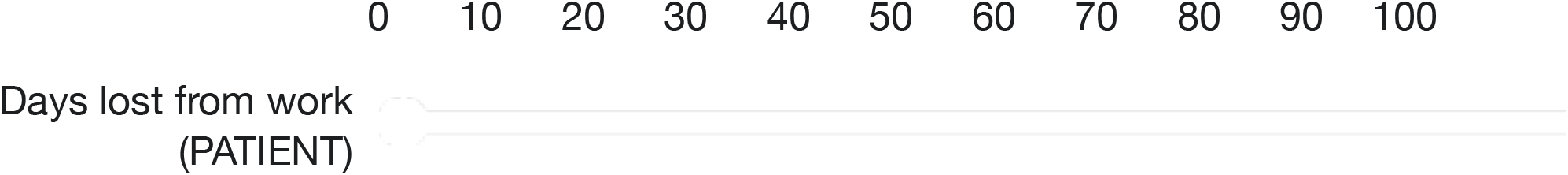

Did a family member miss days of work or school to care for you after this recent illness or injury?

◯ Yes
◯ No

Did the family member who stayed home to care for you WORK or ATTEND SCHOOL prior to your recent illness or injury?

◯ Work (including working at home or family farm)
◯ Attend School
◯ Neither

Please estimate the number of days that ANY FAMILY MEMBER has missed from WORK in order to help care for you after this recent illness or injury

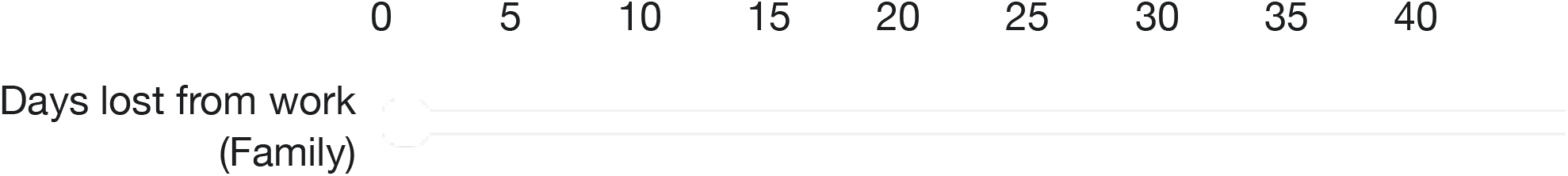

Please indicate what type of work your family member who cared for you engages in

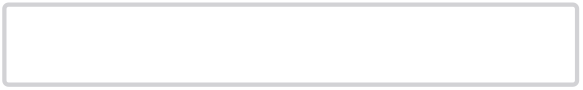

BEFORE YOUR RECENT ILLNESS / INJURY what was FAMILY MEMBER ESTIMATED weekly income?

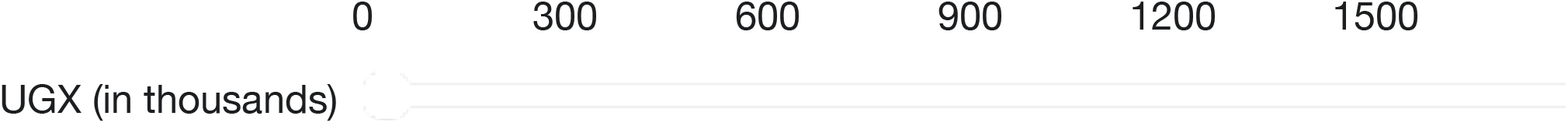

Please estimate the number of days that ANY FAMILY MEMBER has missed from SCHOOL in order to help care for you after this recent illness or injury

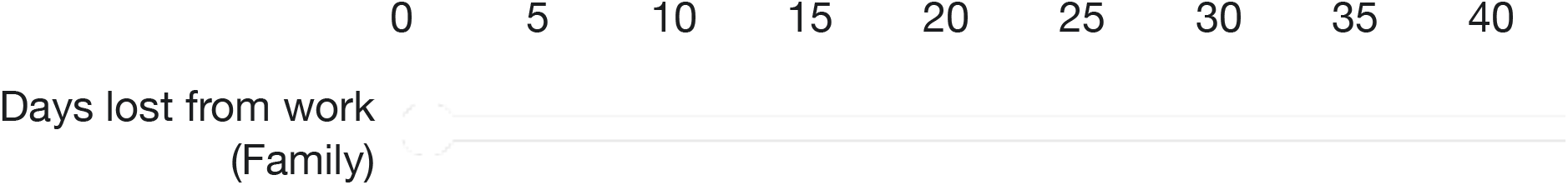

Please estimate your out-of-pocket expenses related to your recent illness or injury (including expense of transport for examination or treatment & cost of any medicines or treatments)

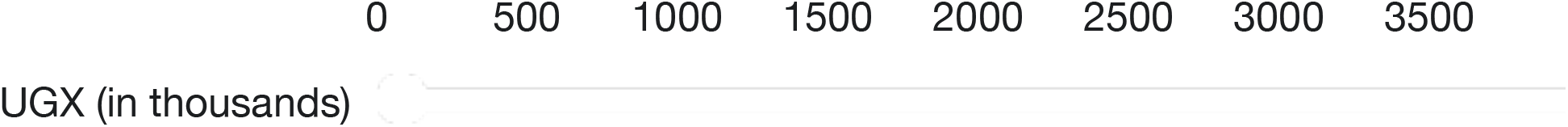

Have you been able to return to WORK after your recent illness or injury?

◯ yes
◯ no

Have you been able to return to SCHOOL after your recent illness or injury?

◯ yes
◯ no

Have you been able to return to USUAL ACTIVITIES after your recent illness or injury?

◯ yes
◯ no

What type of work are you currently doing?

◯ Same work as prior to my illness/injury - full-time
◯ Same work as prior to my illness/injury - part-time
◯ Different work than before my illness/injury
◯ I am not working

Please indicate what type of work you are doing currently

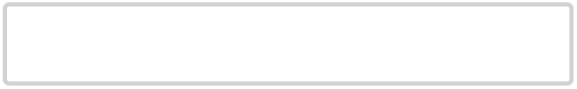

CURRENTLY, what is your estimated weekly income?

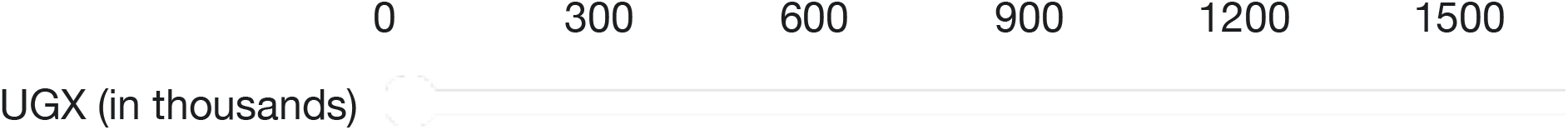

From the time you first became ill/injured, did you have any difficulty paying for any of the following items

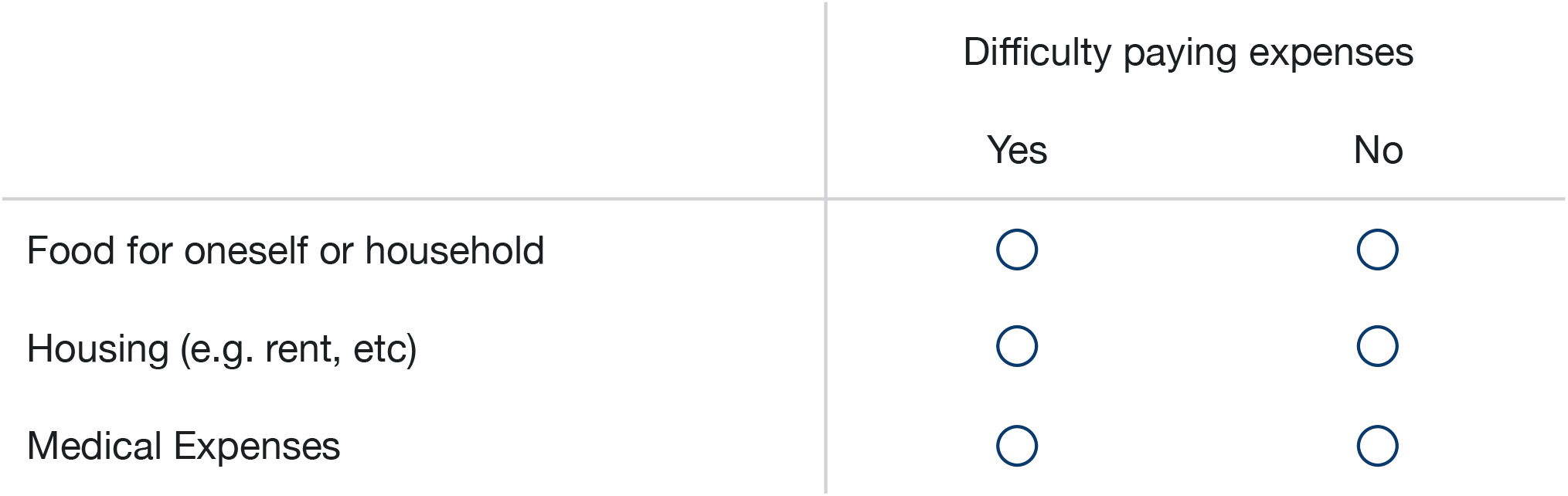

At any time, did your household have to sell any assets to pay for expenses as a result of your recent illness or injury?

◯ Yes
◯ No

Please indicate what asset was sold

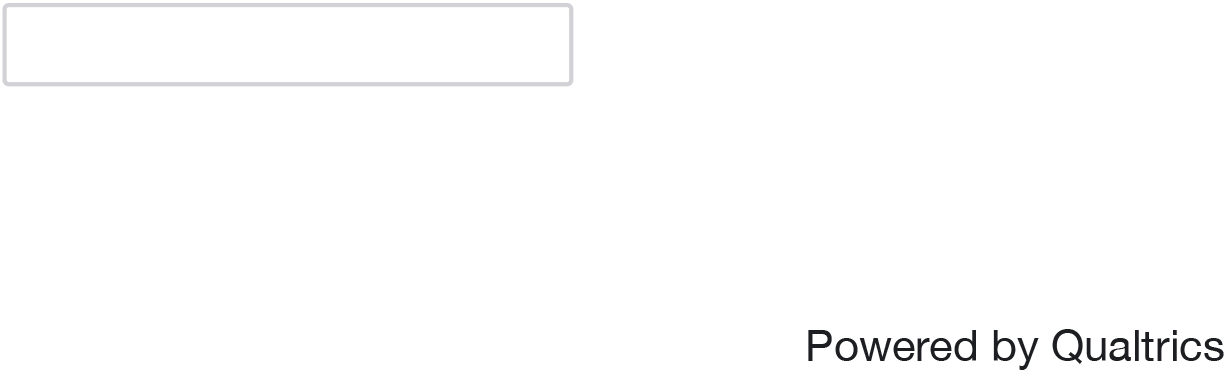

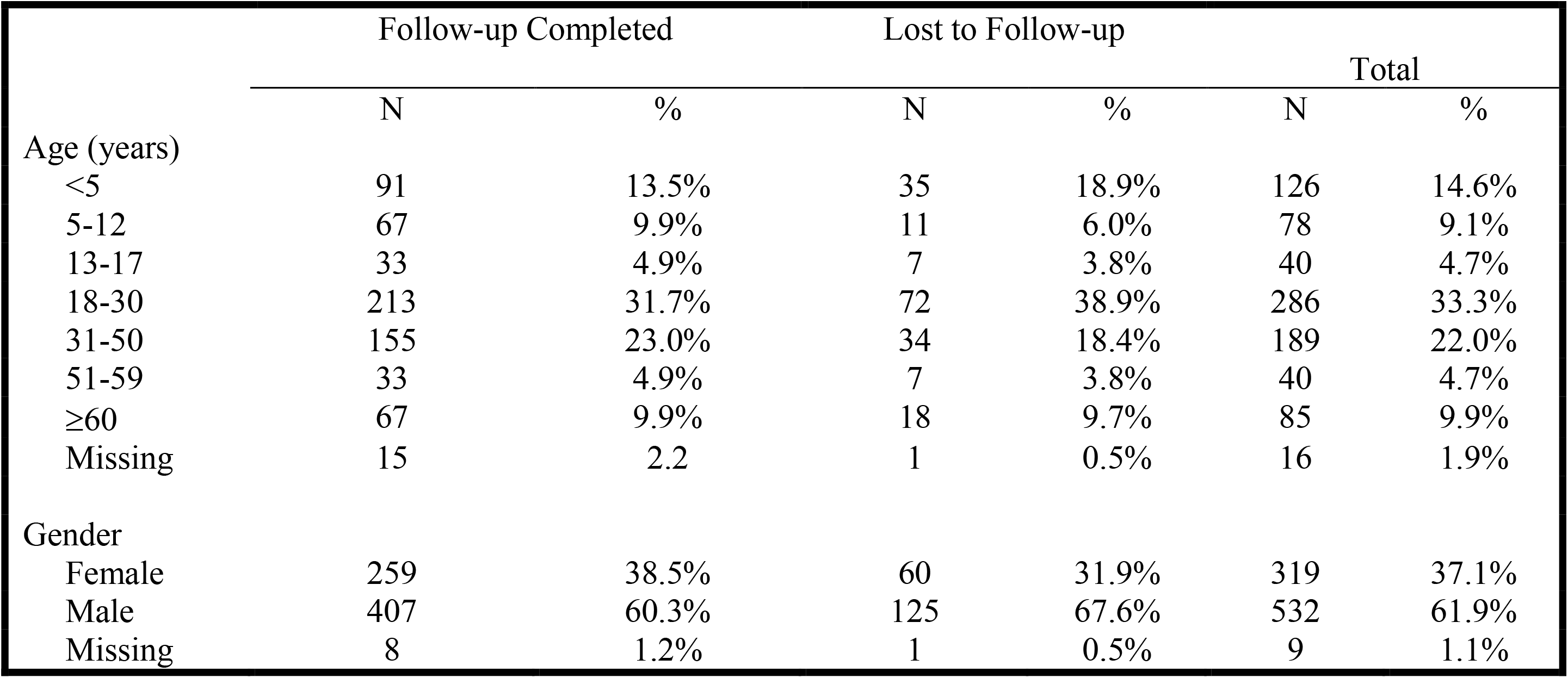

## Appendix 3 Occupation Codebook

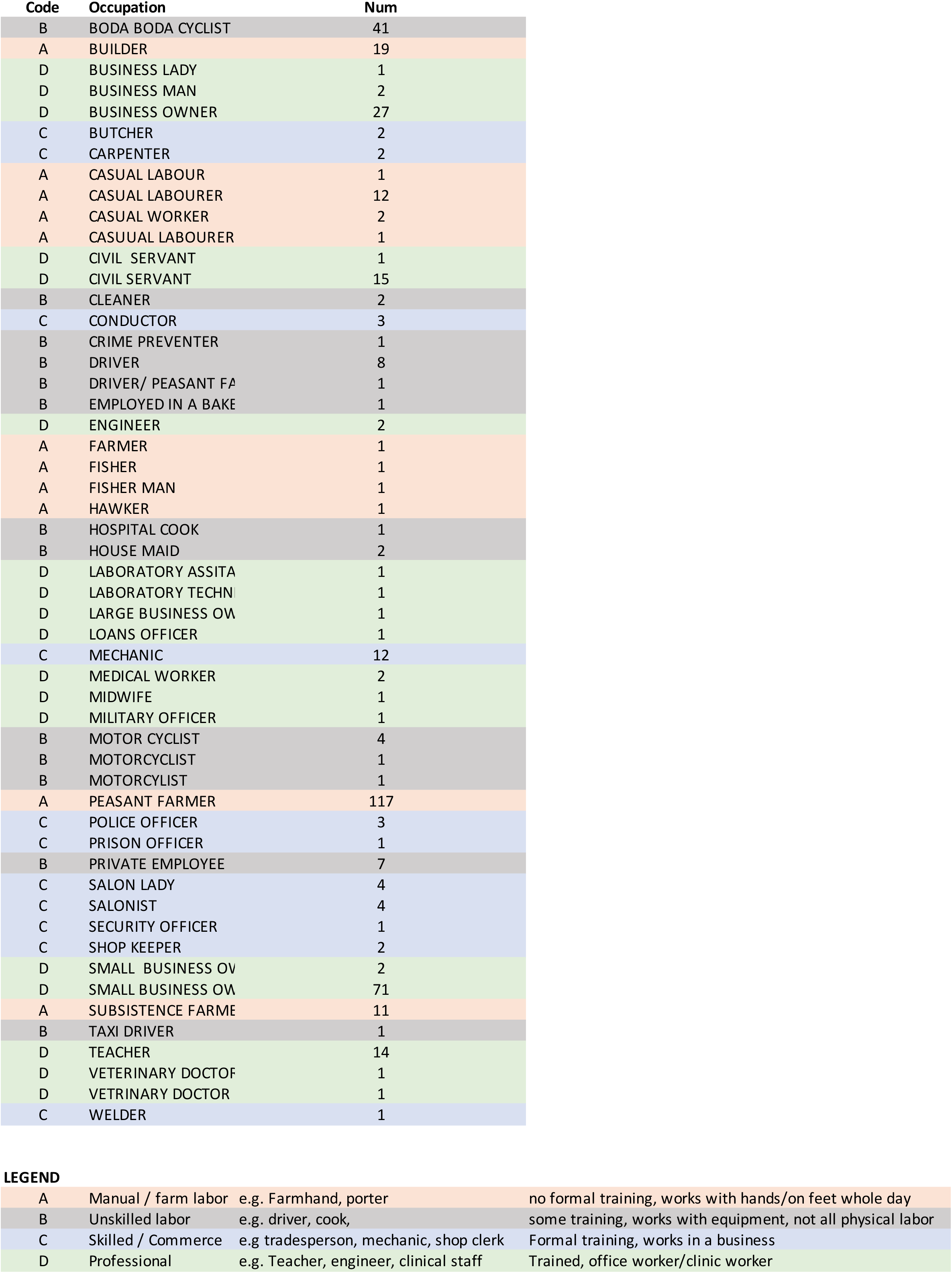

